# Assessment of the infectious threshold of SARS-CoV-2 in primary airway epithelial cells

**DOI:** 10.1101/2021.05.18.21257110

**Authors:** Manel Essaidi-Laziosi, Francisco Javier Perez Rodriguez, Pascale Sattonnet-Roche, Nicolas Hulo, Frederique Jacquerioz, Laurent Kaiser, Isabella Eckerle

## Abstract

Comparison of virus isolation success from clinical samples across a range of viral loads inoculated in parallel on Vero E6 and human airway epithelia (HAE) showed lower success of virus isolation in HAE, suggesting an overestimation of actual infectiousness in humans using Vero E6 cell lines, commonly considered as reference.

## Main text

Understanding the window of Severe Acute Respiratory Syndrome coronavirus 2 (SARS-CoV-2) infectiousness is essential for the control of the coronavirus disease 19 (COVID-19) pandemic. Thanks to its high sensitivity and rapidity, real time RT-PCR remains the gold standard for diagnosing SARS-CoV-2. However, detection of viral RNA by this method does not necessarily equate with the presence of infectious viral particles, the requisite for transmission.

Although resource-intensive (as dependent on biosafety level-3 facilities) and not suitable for routine diagnostic purposes, inoculation of patient’s specimens on cultured cells is to date the only method to confirm presence of infectious SARS-CoV-2 in a sample. A number of studies highlighted two determinants of the presence of infectious particles: SARS-CoV-2 RNA copy numbers (RNAc) per mL in the original specimens, as determined by RT-PCR, and the symptom duration. The probability of isolating viable virus hence appears to be drastically reduced below 5.4-7 log_10_ RNAc/mL, and after more than one week of symptoms [1-3].

The vast majority of these investigations used Vero E6 cells, derived from the kidney of an African Green Monkey, although several other conventional Human cell lines, such as Caco2 (colorectal adenocarcinoma) and Calu3 (lung cancer cells) were also found to be susceptible [4]. As they are interferon-production deficient [5] and express the virus receptor [6], Vero E6 cells are highly susceptible to SARS-CoV-2 and thus are the most widely used reference cell line for isolation. However, these cells do not mimic the *in vivo* situation of the SARS-CoV-2 entry site, which is the human respiratory tract. The use of reconstituted human primary airway epithelial cells (HAE) better reflects SARS-CoV-2 infection characteristics *in vivo*. To assess of the threshold for the presence of infectious virus in this more relevant model system, we compared virus isolation success from clinical samples across a range of viral loads (VLs) inoculated in parallel on Vero E6 and HAE. We found that this threshold is higher in HAE compared to Vero E6, suggesting the presence of infectious virus as determined by Vero E6 would not necessarily be sufficient to lead to an infection in HAE.

In this study, we used nasopharyngeal swabs collected in viral transport medium from symptomatic adult individuals presenting at the outpatient testing center of the University Hospitals of Geneva, spanning the first pandemic wave in spring 2020 and a second pandemic wave in fall 2020, that tested positive for SARS-CoV-2 by RT-PCR (Cobas® SARS-CoV-2 Test, Cobas 6800, Roche, Switzerland). All samples were collected from patients within the first 5 days post onset of symptoms (dpos), diagnosed between April and September 2020. All viruses circulating during the investigated time period were characterized by the D614G mutation, but no variants of concern were circulating in Switzerland during that time. VLs were calculated for the E gene target as previously described [7]. In order to avoid loss of infectivity, all samples were frozen at -80°C after diagnostic testing. Their inoculation was performed immediately after a single thawing (there were no repeated freeze-thaw cycles). Regarding virus isolation, 100 µl of the original clinical sample was inoculated in parallel on Vero E6 cells grown in 48 well plates and in HAE (MucilAir™ commercially available, Epithelix SARL), with an approximate number of cells for both culture systems of 2E+05 cells per well. Infections were performed at 37°C under a 5% CO_2_ atmosphere as previously described [8, 9]. Viral replication was assessed by quantitative RT-PCR from RNA extracted from the supernatant collected at 1 hour post infection (hpi) for Vero E6 and from apical tissue washes 3hpi for HAE, and at the end of the experiment 6 days post infection (dpi). Successful virus isolation was determined by at least 3log10 folds increase of RNAc between baseline and 6 dpi.

In total, 38 primary clinical specimens were inoculated in parallel, with a VL ranging from 5.7-9.0 log_10_ SARS-CoV-2 RNAc/mL in the original sample. The overall frequency of successful virus isolation was 27/38 in Vero E6 and 12/38 in HAE (Figure 1A and Table S1). No growth in either cell culture system was observed in samples below 5.8 log_10_ RNAc/mL. The lowest VL from which infectious virus could be isolated in Vero E6 was 5.9 log_10_ RNAc/mL and 6.3 log_10_ RNAc/mL for HAE (Figures 1A). Although two samples with rather low viral load of 6.3 and 6.5 log_10_ RNAc/mL showed successful isolation in HAE, consistent isolation of virus was only observed in samples with a VL of 7.7 log_10_ RNAc/mL and higher. Using Probit analysis, the probability of a sample being infectious in Vero E6 and HAE was below 5% when VL was lower than 5.5 and 6.5 log_10_ SARS-CoV-2 RNAc/mL, respectively (p value <0.05). No correlation was found between the number of dpos within the first 5 dpos and successful viral growth in both models (in order to increase the likelihood of infectious virus presence, only samples from patients ≤ 5 dpos were selected) (Table S1).

**Figure 1.**
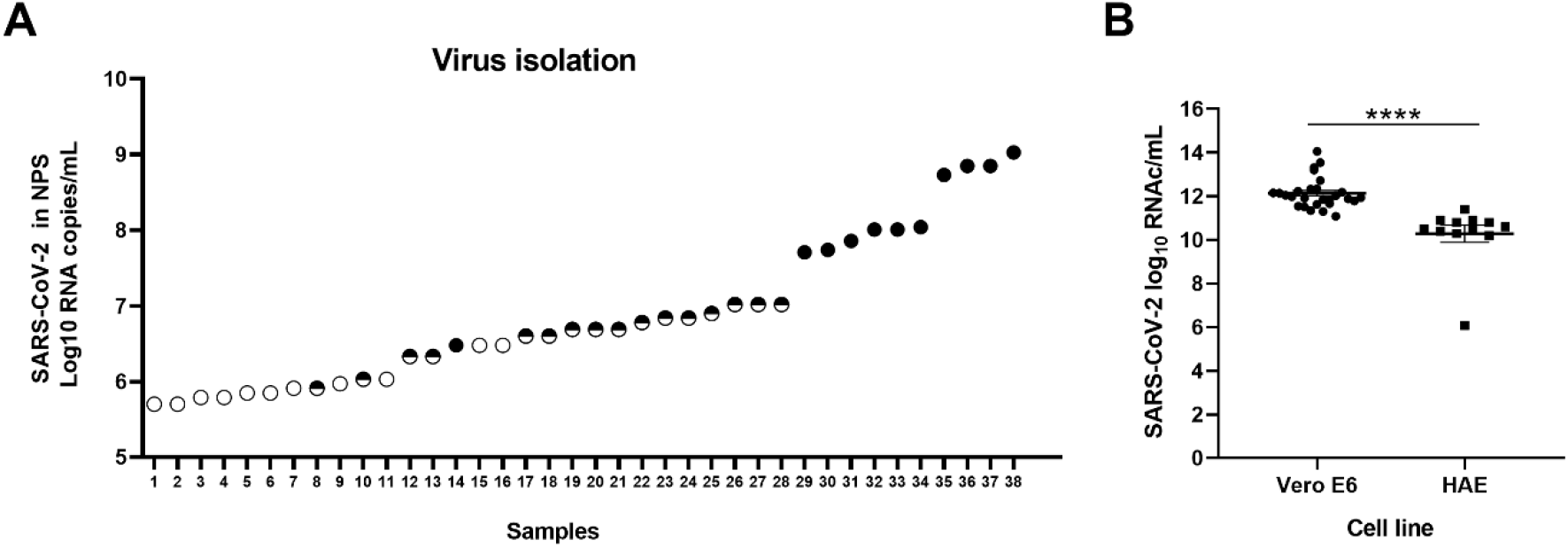
Comparative virus isolation in Vero E6 versus reconstituted human primary airway epithelial cells (HAE). **A** Comparative virus isolation success in Vero E6 versus HAE. Comparing baseline to 6 dpi, the success of isolation was determined and presented as follow: ^○^ : unsuccessful isolation; ◓ : successful isolation only in Vero E6; ● : successful isolation Vero E6 and HAE. **B**. SARS-CoV-2 viral loads 6 dpi after successful isolation in cell culture supernatant (Vero E6) and apical washes (HAE). Statistical significance was calculated using t test (****: p< 0.0001).

Upon successful isolation, the virus showed replication to higher viral loads in Vero E6 (mean VL of 12.1 log_10_ SARS-CoV-2 RNAc/mL, SD ± 0.7, range 11.1-14.06) compared to HAE (mean VL of 10.3 log_10_ SARS-CoV-2 RNAc/mL, SD ± 1.4 range 6.1-11.4) at the end of the experiment (6 dpi, Figure 1B). In conclusion, upon comparative virus isolation, we could recapitulate findings from earlier studies on Vero E6 with successful virus isolation in a similar range as previously reported. In the HAE model, however, virus isolation was only consistently successful in samples with a VLs of ≥7.7 RNAc/mL (Figure 1A), but there were two outliers of successful isolation with a much lower VL. In conclusion, Vero E6 cells are shown to be more permissive for SARS-CoV-2 isolation and yield virus isolates from samples with lower VLs than HAE. Furthermore, upon successful isolation, SARS-CoV-2 replicates in Vero E6 to higher virus titers than in HAE.

Lower success of virus isolation in HAE, and thus a higher amount of infectious virus particles needed to start an infection, could hint towards an overestimation of actual infectiousness in humans when only using Vero E6 cell lines as a reference. This could mean that the actual transmission risk would be lower than findings based on Vero E6 culture data and that transmission-relevant infectious virus shedding is shorter and/or lower than what was previously estimated. Furthermore, antigen-based rapid diagnostic tests, which have a lower sensitivity than PCR but may detect infections with VLs as low as 6 log_10_ RNAc/mL [10], would thus be an even better at identifying infectious individuals than previously believed. Further research is still needed to investigate the infectious doses of SARS-CoV-2 under real life conditions. Even if the presence of such infectious virus is not enough to predict the transmission risk, as multiple factors influence transmission, VL constitutes one of the most important drivers of transmission [11].

As the infectious dose of SARS-CoV-2 still needs to be determined *in vivo* [12], and HAEs do not completely recapitulate the *in vivo* situation, our study cannot be used to change existing guidelines on isolation or discharge criteria. It nevertheless emphasizes the effect of the cell lines used to culture the virus and thus supports the use of models that more closely reflect the *in vivo* situation, such as HAE in air-liquid interface culture rather than conventional cell lines, in order to better understand transmission risks of SARS-CoV-2 in patients.

## Data Availability

The authors confirm that the data supporting the findings of this study are available within the article and its supplementary materials.

## Acknowledgments

We thank Catia Alvarez for excellent technical support and Erik Boehm for English proof reading (Geneva Centre for Emerging Viral Diseases, Geneva University Hospitals, Geneva, Switzerland).

---

**Table S1.**
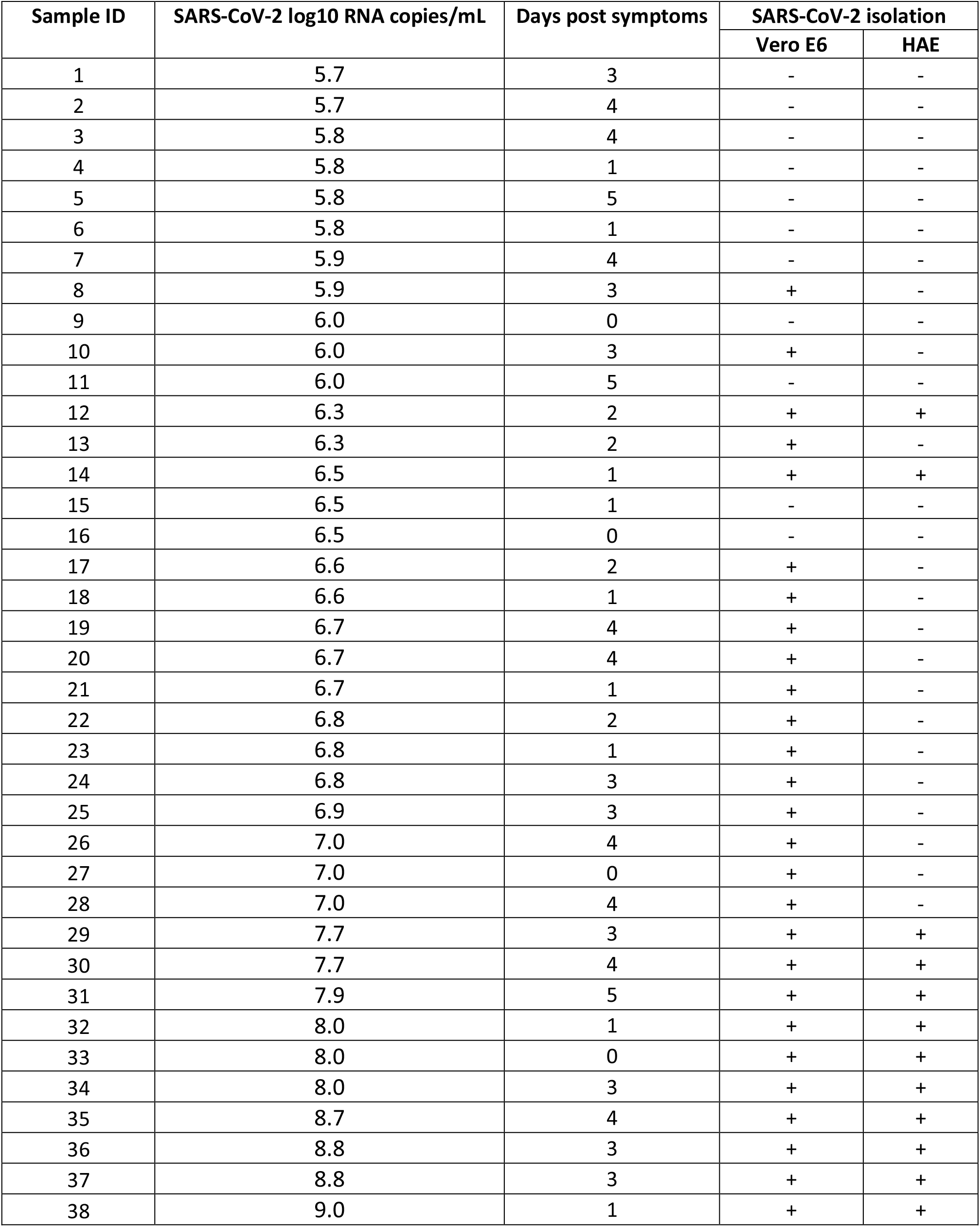
Sample characteristics

## References

1. Wölfel R, Corman VM, Guggemos W, et al. Virological assessment of hospitalized patients with COVID-2019. Nature 2020; 581(7809): 465–9.

2. Bullard J, Dust K, Funk D, et al. May 2020. Predicting infectious SARS-CoV-2 from diagnostic samples. Clin Infect Dis doi 10.

3. van Kampen JJ, van de Vijver DA, Fraaij PL, et al. Duration and key determinants of infectious virus shedding in hospitalized patients with coronavirus disease-2019 (COVID-19). Nature communications 2021; 12(1): 1–6.

4. Chu H, Chan JF-W, Yuen TT-T, et al. Comparative tropism, replication kinetics, and cell damage profiling of SARS-CoV-2 and SARS-CoV with implications for clinical manifestations, transmissibility, and laboratory studies of COVID-19: an observational study. The Lancet Microbe 2020; 1(1): e14–e23.

5. Chew T, Noyce R, Collins SE, Hancock MH, Mossman KL. Characterization of the interferon regulatory factor 3-mediated antiviral response in a cell line deficient for IFN production. Molecular immunology 2009; 46(3): 393–9.

6. Ren X, Glende J, Al-Falah M, et al. Analysis of ACE2 in polarized epithelial cells: surface expression and function as receptor for severe acute respiratory syndrome-associated coronavirus. Journal of general virology 2006; 87(6): 1691–5.

7. Baggio S, L’Huillier AG, Yerly S, et al. Severe Acute Respiratory Syndrome Coronavirus 2 (SARS-CoV-2) Viral Load in the Upper Respiratory Tract of Children and Adults With Early Acute Coronavirus Disease 2019 (COVID-19). Clinical Infectious Diseases 2020.

8. L’Huillier AG, Torriani G, Pigny F, Kaiser L, Eckerle I. Culture-competent SARS-CoV-2 in nasopharynx of symptomatic neonates, children, and adolescents. Emerging infectious diseases 2020; 26(10): 2494.

9. Essaidi-Laziosi M, Brito F, Benaoudia S, et al. Propagation of respiratory viruses in human airway epithelia reveals persistent virus-specific signatures. Journal of Allergy and Clinical Immunology 2018; 141(6): 2074–84.

10. Berger A, Nsoga MTN, Perez-Rodriguez FJ, et al. Diagnostic accuracy of two commercial SARS-CoV-2 Antigen-detecting rapid tests at the point of care in community-based testing centers. Plos one 2021; 16(3): e0248921.

11. Marks M, Millat-Martinez P, Ouchi D, et al. Transmission of COVID-19 in 282 clusters in Catalonia, Spain: a cohort study. The Lancet Infectious Diseases 2021.

12. Karimzadeh S, Bhopal R, Tien HN. Review of infective dose, routes of transmission, and outcome of COVID-19 caused by the SARS-CoV-2 Virus: comparison with other respiratory viruses. 2020.

