## Supplemental table1 for "Assessment of the infectious threshold of SARS-CoV-2 in primary airway epithelial cells": Essaidi-Laziosi MedRxiv virus isolatio_supp data.pdf

| Sample ID | SARS-CoV-2 log10 RNA copies/mL | Days post symptoms | SARS-CoV-2 isolation | |
|---|---|---|---|---|
| | | | Vero E6 | HAE |
| 1 | 5.7 | 3 | - | - |
| 2 | 5.7 | 4 | - | - |
| 3 | 5.8 | 4 | - | - |
| 4 | 5.8 | 1 | - | - |
| 5 | 5.8 | 5 | - | - |
| 6 | 5.8 | 1 | - | - |
| 7 | 5.9 | 4 | - | - |
| 8 | 5.9 | 3 | + | - |
| 9 | 6.0 | 0 | - | - |
| 10 | 6.0 | 3 | + | - |
| 11 | 6.0 | 5 | - | - |
| 12 | 6.3 | 2 | + | + |
| 13 | 6.3 | 2 | + | - |
| 14 | 6.5 | 1 | + | + |
| 15 | 6.5 | 1 | - | - |
| 16 | 6.5 | 0 | - | - |
| 17 | 6.6 | 2 | + | - |
| 18 | 6.6 | 1 | + | - |
| 19 | 6.7 | 4 | + | - |
| 20 | 6.7 | 4 | + | - |
| 21 | 6.7 | 1 | + | - |
| 22 | 6.8 | 2 | + | - |
| 23 | 6.8 | 1 | + | - |
| 24 | 6.8 | 3 | + | - |
| 25 | 6.9 | 3 | + | - |
| 26 | 7.0 | 4 | + | - |
| 27 | 7.0 | 0 | + | - |
| 28 | 7.0 | 4 | + | - |
| 29 | 7.7 | 3 | + | + |
| 30 | 7.7 | 4 | + | + |
| 31 | 7.9 | 5 | + | + |
| 32 | 8.0 | 1 | + | + |
| 33 | 8.0 | 0 | + | + |
| 34 | 8.0 | 3 | + | + |
| 35 | 8.7 | 4 | + | + |
| 36 | 8.8 | 3 | + | + |
| 37 | 8.8 | 3 | + | + |
| 38 | 9.0 | 1 | + | + |

Table S1. Sample characteristics
